# Administration Technique of Intranasal Corticosteroid Sprays among Nepali Pharmacists: A Cross-Sectional Study

**DOI:** 10.1101/2025.06.29.25330472

**Authors:** Amar Prashad Chaudhary, Suraj Thakur, Shiv Kumar Sah

**Affiliations:** Department of Pharmacy Practice, Institute of Medicine, Tribhuvan University, Kathmandu, Nepal

**Keywords:** intranasal corticosteroid spray, allergic rhinitis, device use technique, pharmacist, Patient Counselling, Continuing Pharmacy Education

## Abstract

**Background:** Allergic rhinitis is a common condition affecting up to 40% of people worldwide, with a notably high prevalence in South Asia. The primary treatment for moderate to severe allergic rhinitis is intranasal corticosteroid sprays, typically demonstrated to patients by registered pharmacists. However, many patients do not use these sprays correctly. This study evaluated the proficiency of pharmacists in demonstrating the correct technique for using intranasal corticosteroid sprays and the factors contributing to proper technique.

**Methods:** In a cross-sectional survey of 365 registered pharmacists in the Kathmandu Valley, Nepal, a trained observer utilized a standardized 12-step checklist to assess each pharmacist’s technique for using intranasal corticosteroid sprays. Demographics, education, experience, previous training, and instructional materials use were recorded. Proficiency was classified as “adequate” if more than 6 out of 12 marks were obtained.

**Results:** Out of 365 pharmacists, 65.5% were male, and approximately 59% were aged 26 or younger. About 70% of pharmacists hold a diploma in pharmacy. More than half of the pharmacists demonstrated inadequate technique, while 47.1% showed adequate skill overall. However, only 6% could demonstrate all 5 critical steps. The likelihood of providing appropriate counselling on the use of intranasal corticosteroid sprays was significantly correlated with male gender, older age, use of educational materials, conducting moderate counselling sessions weekly, and higher qualifications.

**Conclusions:** More than half of the registered pharmacists in Nepal demonstrated inadequate technique of intranasal corticosteroid sprays. There is a strong need for educational intervention and policy change for improved proficiency.

## Background

A chronic inflammatory condition of the nasal mucosa, allergic rhinitis (AR) is brought on by immunoglobulin E (IgE) mediated responses to allergens breathed. There are many causes of allergic rhinitis, including Pollen, dust mites, Cockroach waste, animal dander, fumes and odors, changes in environment, smoke, and certain foods or spices. The most common symptoms of AR are sneezing, stuffy nose, runny nose, Itchy nose, throat, eyes, and ears, nosebleeds, clear drainage from the nose, snoring, and breathing through the mouth.

AR affects 10% to 40% of the world’s population, and its prevalence is increasing in many countries [1,2]. AR and other allergy disorders are also common in Nepal and the surrounding South Asian nations. A recent school-based study in Nepal, for example, found rhino conjunctivitis symptoms in 28% of children [3]. AR was responsible for almost 25% of allergy illnesses in Nepal’s Gandaki Province [3]. Adolescent AR prevalence in India is estimated at 22%, whereas in adults it was found to be 11% and 33.3% in asthmatics [4,5]. Similarly, a large-scale study conducted in Europe discovered that up to 20% of the population is impacted by AR [6]. The prevalence of allergic rhinitis in the United States of America is slightly lower, is 7.7% in adults and 7.2% in children [7].

Therefore, the treatment of AR is very important as it impacts daily life activities. The objective of AR treatment is to control the disease. Antihistamines, leukotriene receptor antagonists, azelastine, and intranasal corticosteroid spray (INCS) are used for treating AR according to the Allergic Rhinitis and its Impact on Asthma (ARIA) guidelines 2019 [8]. Effective pharmacotherapy is crucial for symptomatic control of AR. INCS sprays are the most potent medications for moderate to severe AR and are recommended as first-line therapy [9]. When used correctly, INCS reduce nasal congestion, rhinorrhea, sneezing, and itching by suppressing mucosal inflammation.

The most common adverse drug reactions of INCS include dyspnea, anosmia, ageusia/dysgeusia, epistaxis, and headache [10]. A study conducted at the ENT outpatient clinic at Aberdeen Royal Infirmary found that 15.5% reported epistaxis due to an ipsilateral hand technique [11]. Similarly, a study in Thailand discovered a 3.6 times higher risk of adverse events in patients who did not point the tip of the spray away from the nasal septum [12]. Maintaining a neutral head position and exhaling through the mouth are crucial for proper drug disposition and enhanced efficacy [13]. Therefore, using the correct technique is vital for better efficacy and a reduced risk of side effects. Standard guidelines recommend instructing patients to shake the spray, maintain a neutral head position, insert the tip slightly upward and laterally (away from the septum), close the opposite nostril, inhale gently while actuating the spray, and then exhale through the mouth [12,14].

However, a study conducted by Rattanawang et al. found that only 4% of patients performed all twelve steps, while only 29% completed all the crucial steps [12]. Similarly, a study by Gurung et al. in Nepal revealed that only 7.2% of patients executed all the steps correctly, and 18.2% managed to perform all five critical steps accurately [15]. A systematic review indicated that approximately 73% of patients did not receive proper advice regarding nasal corticosteroid sprays [16].

Healthcare professionals, especially pharmacists, are responsible for counselling the patient regarding the drugs dispensed by them. Given this context, it is essential to assess how well Nepali registered pharmacists themselves understand and can demonstrate correct INCS technique. No prior studies have examined this. By identifying gaps in pharmacist knowledge and technique, targeted interventions (e.g., curriculum changes or training modules) can be designed to improve AR care. This study therefore evaluated the proficiency of registered pharmacists in Kathmandu Valley in demonstrating INCS administration, and analyzed professional factors associated with adequate technique. The findings have important implications for Nepal’s healthcare and pharmacy education systems: improving pharmacist competency could raise the standard of AR management nationally.

## Methods

### Study design and Study period

A cross-sectional observational study was performed from 1 November 2023 to 28 May 2024 through interviews of registered pharmacists. They answered a semi-structured questionnaire containing questions about their sociodemographic, professional details, and INCS counselling steps. STROBE (Strengthening the Reporting of Observational Studies in Epidemiology) principles were adhered to in the study’s reporting [17,18].

### Study population and study site

The sample was selected from pharmacists registered at the Nepal Pharmacy Council working at community pharmacies in Kathmandu, Nepal. Being Nepal’s capital, Kathmandu is a heavily populated city. The respective site had a large number of community pharmacies, about 4000, with many registered pharmacists [19].

### Sampling method

Simple random sampling of the community pharmacies available in different wards of Kathmandu district of Nepal was done. This cross-sectional study identified potential participants from the registered pharmacists working at community pharmacies. These potential participants were approached and explained the study’s purpose, procedures, and potential risks and benefits. The same interviewer interviewed all the participants to overcome the interobserver variability in participants’ responses.

### Sample size

The survey study was completed using the Raosoft sample size calculator to capture the appropriate sample size [20]. A minimum of 363 samples was required for a 95% confidence interval and a 5% margin of error for the population distribution of 21000 registered pharmacists at a 40% response distribution [21]. Thus, a total of 365 registered pharmacists participated in this study.

### Measures

After the individuals’ sociodemographic and professional information was obtained through interviews, the 12-step nasal spray application technique as given in Table 1 was demonstrated by the participant and was examined [12,13,22–24]. Each correct step was assigned 1 mark, and the incorrect or missed step was assigned 0 marks. Hence, the maximum score obtained was 12 marks. Five steps in INCS counselling which contains asterisk in the Table 1 were considered critical steps that impact the patient outcome and risk of adverse drug reactions. Based on the study conducted by Binita et al., expert suggestions, and the median value, 6 marks was established as the cut-off score [25]. The median value of the total marks scored was 6. Consequently, anyone with a score higher than 6 marks was categorized as performing adequately, and anyone with marks equal to or lower than 6 was categorized as performing inadequately.

**Table 1:** Steps for the administration of the INCS spray.

| S. N | Steps for the administration of the INCS spray |
| --- | --- |
| 1. | Shake the spray in a vertical plane |
| 2. | Remove the dust cap |
| 3. | Blow the nose* |
| 4. | Hold the spray bottle, pointing the tip of the nozzle up with the hand |
| 5. | Replace the index and middle finger on the pusher and the thumb at the bottom of a spray bottle |
| 6. | Put the tip of the nozzle into the nostril and close the other side |
| 7. | Neutral head or slightly tilt the head forward* |
| 8. | Point the tip slightly outwards, away from the septum* |
| 9. | Squirt the spray in the nose while breathing in* |
| 10. | Breathe out through the mouth* |
| 11. | Wipe the nozzle with a tissue or handkerchief |
| 12. | Replace the cap |

*\*=critical steps*

### Reliability and Validity

The initial questionnaire was validated by a subject expert, composed of advisors, professors, and teachers, for correctness, clarity, appropriateness, and jargon use. This validation was conducted using face validity approaches. An inter-rater reliability test was conducted on 15 participants and found a Cronbach’s alpha value of 0.758.

### Inclusion and Exclusion Criteria

This study only took into account pharmacists aged 18 years and above who were registered with the Nepal Pharmacy Council and employed in community pharmacies. Specifically, those with a Diploma in Pharmacy (DPharm) degree, Bachelor in Pharmacy (BPharm) degree, Doctor in Pharmacy (PharmD) degree, and Master in Pharmacy (MPharm) degree. Participants should have a minimum of one year of experience. No unregistered pharmacists, Pharmacy students, or Interns were considered for this study.

### Data Collection Procedure

The essential information was then gathered from participants using a semi-structured questionnaire through in-person interview. The standardized protocol was followed during an interview. Prior to their enrolment in the study, all participants were informed of its purpose, and their consent was acquired.

### Statistical analysis

Utilizing Microsoft Excel and the Statistical Package for Social Sciences (SPSS) version 26 software, the gathered data was analyzed. Factors related to the administration technique were evaluated using multivariate binary logistic regression to understand their independent impact. The decision tree analysis was done using Chi square Automatic Interaction Detector (CHAID) to compliment the findings of univariate logistic regression. When the P-value was less than 0.05 and the confidence level was 95%, it was deemed statistically significant.

### Ethical Considerations

Ethical approval reference no. 210 (6-11) E2, 080/081 was taken from the Institutional Review Committee before the commencement of the study. Informed consent was taken from the participant before any data were collected from the study site.

## Results

### Participant Characteristics

Pharmacists’ professional and demographic traits are listed in Table 2. The study involved 365 registered pharmacists as participants. Most were under 26 years old (59.2%), and most were men (65.5%). Approximately two-thirds of those who took part were single. Only roughly one-third of participants (30%) had a Bachelor of Pharmacy degree or above, whereas the majority (70%) had a Diploma in Pharmacy degree. Moreover, over three-fourths of the participants had early careers (1-4 years), whereas about one-fourth had mid or late pharmaceutical careers (5 years and above).

**Table 2.** Demographic and professional characteristics of pharmacists (N=365).

| **Variables** | **Frequency** | **Percentage** |
| --- | --- | --- |
| **Sex** |  |  |
| Male | 239 | 65.5 |
| Female | 126 | 34.5 |
| **Age** |  |  |
| Less than and equal to 26 | 216 | 59.2 |
| More than and equal to 27 | 149 | 40.8 |
| **Marital status** |  |  |
| Unmarried | 244 | 66.8 |
| Married | 121 | 33.2 |
| **Qualification** |  |  |
| D pharm | 255 | 69.9 |
| B pharm and above | 110 | 30.1 |
| **Years of experience** |  |  |
| 1-4 years | 267 | 73.2 |
| 5 years and above | 98 | 26.8 |
| **INCS counseling (Weekly)** |  |  |
| Occasionally | 119 | 32.6 |
| 1-4 times | 194 | 53.2 |
| More than 4 times | 52 | 14.2 |
| **Received training** |  |  |
| Yes | 30 | 8.2 |
| No | 335 | 91.8 |
| **Use of information material** |  |  |
| Yes | 75 | 20.5 |
| No | 290 | 79.5 |

More than half have counselled patients on intranasal corticosteroids one to four times a week, but only 8.2% acknowledged any formal training in INCS administration. Additionally, just 20.5% of participants used informative resources.

### Administration Technique Adherence and Proficiency Level

Among 365 participating pharmacists, adherence to INCS administration steps varied widely, as shown in Table 3. High adherence (>80%) was observed in four basic steps: removing the dust cap, replacing the cap, shaking the spray, and holding the bottle upright. In addition, moderate adherence (40–80%) was noted in three steps involving inhaling while spraying, finger positioning, and nozzle insertion. However, low adherence (<40%) was seen in five steps, among which four were critical: blowing the nose, pointing the nozzle away from the septum, exhaling through the mouth, proper head positioning, and wiping the nozzle post-use.

**Table 3:** Performance of each administration step by pharmacists (N=365).

| S. N | Steps for the administration of the INCS spray | Frequency | Percentage |
| --- | --- | --- | --- |
| 1. | Shake the spray in a vertical plane | 309 | 84.7 |
| 2. | Remove the dust cap | 365 | 100 |
| 3. | Blow the nose* | 39 | 10.7 |
| 4. | Hold the spray bottle, pointing the tip of the nozzle up with the hand | 293 | 80.3 |
| 5. | Replace the index and middle finger on the pusher and the thumb at the bottom of a spray bottle | 220 | 60.3 |
| 6. | Put the tip of the nozzle into the nostril and close the other side | 146 | 40 |
| 7. | Neutral head or slightly tilt the head forward* | 122 | 33.4 |
| 8. | Point the tip slightly outwards, away from the septum* | 36 | 9.9 |
| 9. | Squirt the spray in the nose while breathing in* | 287 | 78.6 |
| 10. | Breathe out through the mouth* | 43 | 11.8 |
| 11. | Wipe the nozzle with a tissue or handkerchief | 123 | 33.7 |
| 12. | Replace the cap | 359 | 98.4 |

The participants’ median score across all marks was 6. However, the five crucial steps only had a median score of 1. Twelve points were awarded for INCS counselling, and five points were awarded for the five critical steps. Just 6% of participants were able to accurately complete all five critical steps. More than half of the registered pharmacists were found to be inadequately knowledgeable on INCS patient counselling. Only 46.3% of respondents knew enough about INCS counselling, as seen in Figure 1.

**Fig 1:**
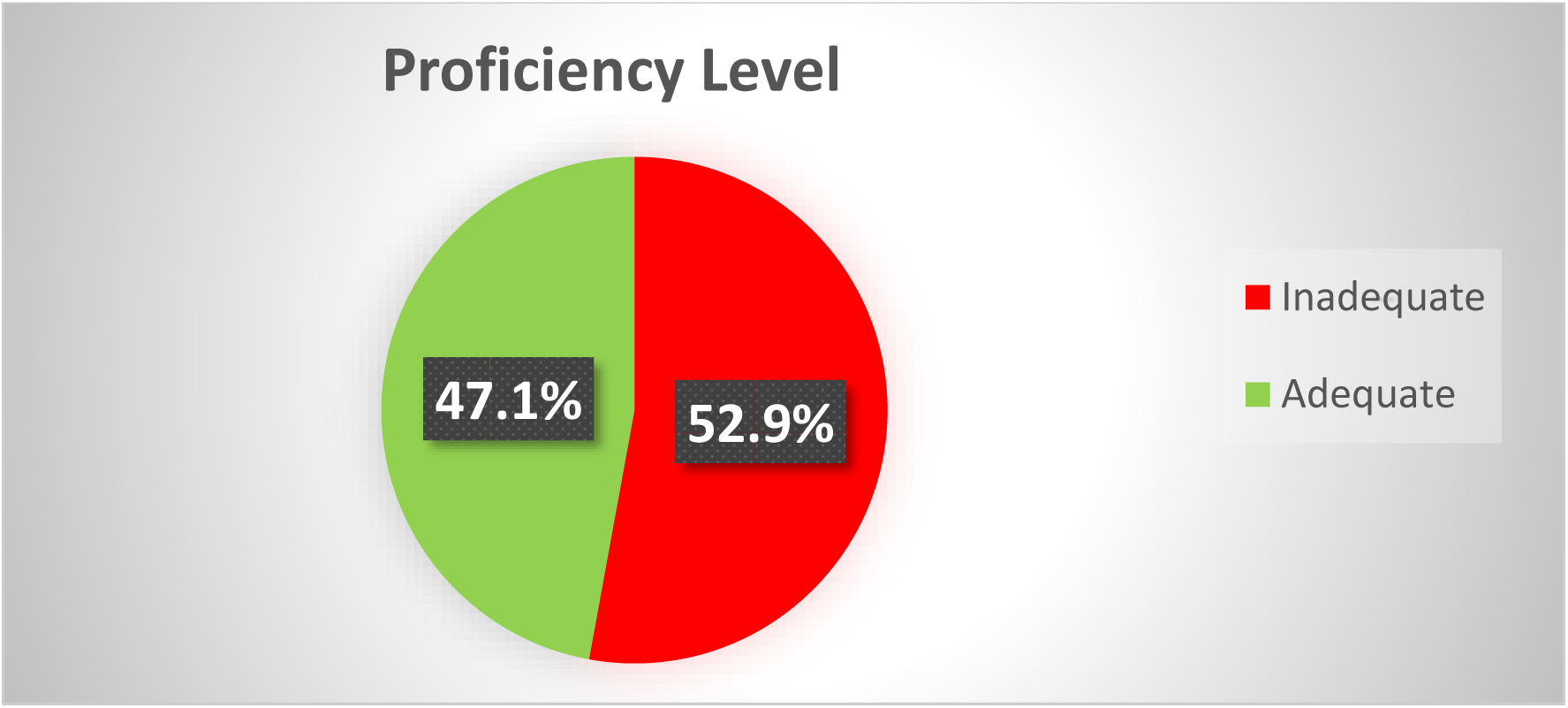
Proficiency level of registered pharmacists regarding the proper administration technique of INCS sprays

### Factors associated with proper administration technique

Several professional and sociodemographic factors were shown to be substantially correlated with the degree of administration technique proficiency by the multivariate binary logistic regression analysis (Table 4). Years of experience, training and marital status did not show statistically significant relationships, while sex, age, qualification, frequency of patient counselling weekly, and the utilization of information material were found to be significant predictors.

**Table 4:** Binary logistic regression analysis of proficiency level of administration technique and different sociodemographic and professional details variables.

| Variable | aOR | CI | P-value |
| --- | --- | --- | --- |
| Sex |  |  |  |
| Male | 2.30 | 1.11 – 4.75 | 0.025 |
| Female | Ref |  |  |
| Age |  |  |  |
| More than 26 years | 0.11 | 0.03 – 0.41 | 0.001 |
| less than or equal to 26 years | Ref |  |  |
| Marital Status |  |  |  |
| Married | 2.39 | 0.71 – 8.06 | 0.160 |
| Unmarried | Ref |  |  |
| Training |  |  |  |
| Yes | - | - | 0.998 |
| No | Ref |  |  |
| Use of Information Material |  |  |  |
| Yes | 0.04 | 0.004 – 0.38 | 0.005 |
| No | Ref |  |  |
| Qualification |  |  |  |
| BPharm and above | 0.03 | 0.007 – 0.14 | 0.000 |
| Dpharm | Ref |  |  |
| years of experience |  |  |  |
| 5 years and above | 0.80 | 0.33 – 1.94 | 0.625 |
| 1-4 years | Ref |  |  |
| INCS counseling (weekly) |  |  |  |
| 1- 4 times | 4.80 | 0.91 – 25.30 | 0.064 |
| Above 4 times | 11.21 | 2.35 – 53.53 | 0.002 |
| Occasionally | Ref |  |  |

The likelihood of men pharmacists exhibiting proper technique was about two times higher than that of female pharmacists. The probabilities of using an inappropriate INCS counselling technique were 89% lower for individuals who were older than 26 years. Proficiency was substantially predicted by having used educational materials. Pharmacists who used educational materials had a 96% less likely to perform inadequately. Pharmacists with a BPharm degree or higher were also around 97% less likely to counsel inappropriately than those with a DPharm degree.

According to this study, individuals who advise patients on INCS more than four times per week were 11 times more likely to demonstrate proficiency as opposed to those who counsel occasionally.

The classification tree (CHAID method), as shown in Figure 2, was developed to identify key predictors of pharmacist proficiency in INCS counselling. The final pruned classification included five levels with nine terminal nodes, achieving an overall classification accuracy of 81.6%.

**Fig 2:**
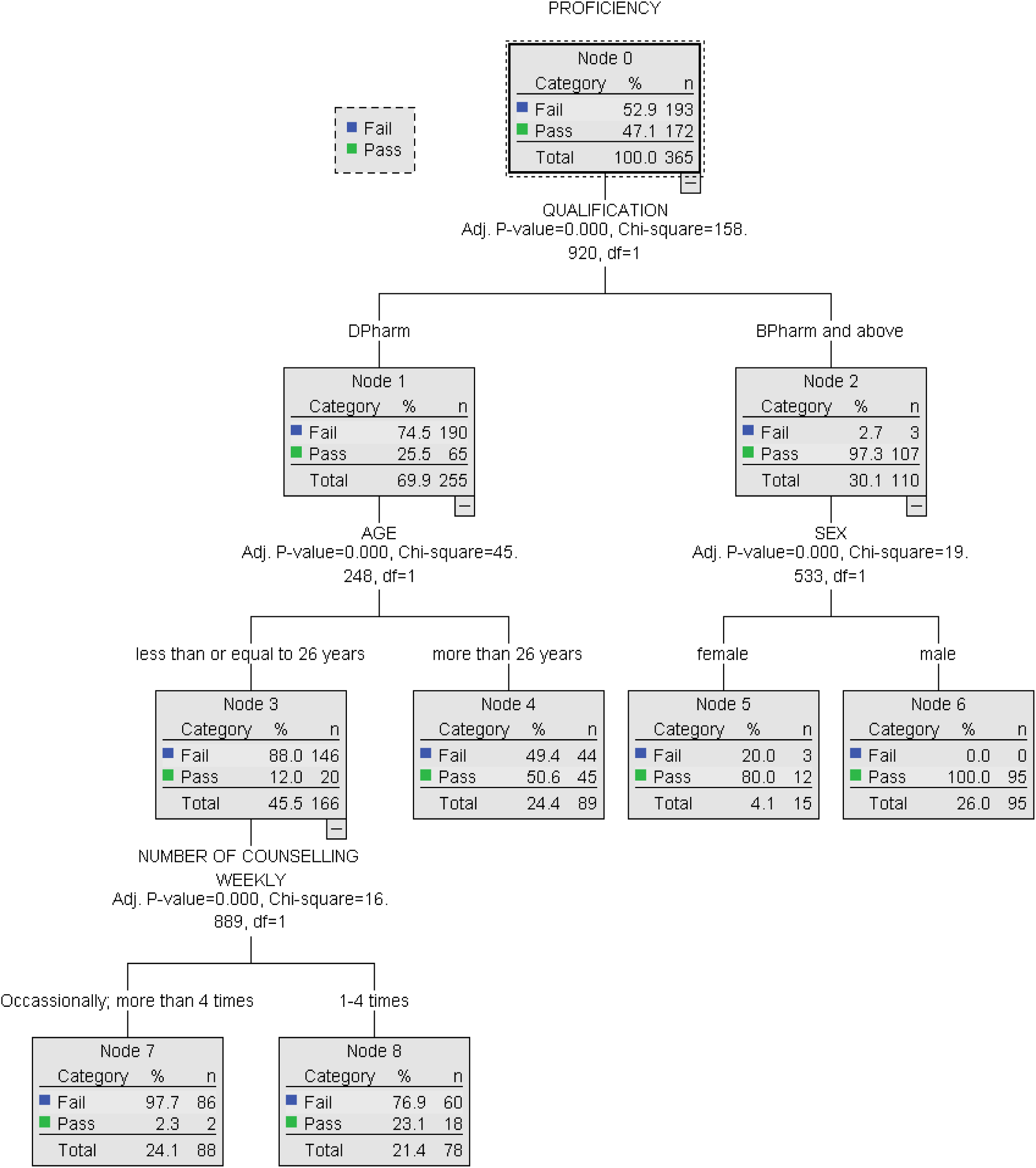
**Classification Tree model of predictors for proficient INCS counselling among pharmacists.**

The first and the most significant split was based on the educational qualification. The pharmacist with a DPharm degree had less proficiency (Fail: 74.5%). However, pharmacists with a BPharm degree or higher had a high proficiency rate (Pass: 97.3%). Among the Dpharm group, age was another significant predictor. Those aged less than 26 years had a pass rate of 12%, whereas the older peers had a pass rate of 50. Similarly, among the Bpharm group, another major factor was gender. Male pharmacists were found to be 100% proficient in INCS counseling, whereas females were found to be only 80% proficient.

Finally, for older Dpharm degree holders (≥ 26 years old) frequency of INCS counselling was another predictor. Those older participants who counseled occasionally or more than four times a week had significantly low proficiency (Fail: 97.7%) compared to those who counseled 1 to 4 times weekly (Fail: 76.9%).

## Discussion

This study addresses a critical gap in pharmacist competency regarding INCS within resource-constrained health systems, where pharmacists are frontline care providers. This research is one of a kind, conducted in Nepal. The survey revealed a significant gap in the participants’ understanding of INCS counseling which helps to understand its impact on the health outcomes of the patient. Approximately 50% of the pharmacists lacked adequate INCS counseling abilities. According to this study, only 6% of pharmacists were able to complete all the essential patient counseling steps, which are crucial for appropriate drug disposal and minimizing the risk of adverse drug reactions. Classification tree analysis showed that educational degree was the primary predictor for the INCS counselling proficiency. BPharm degree or higher degree holders were far more proficient than DPharm degree holders.

The survey’s conclusions about the inadequate INCS administration abilities of Nepali registered pharmacists are in line with the findings of patients and medical professionals worldwide [2,14,26]. And only 6% of pharmacists performed all recommended steps correctly, which was also similar to the study of healthcare workers in Thailand [14]. However, even in a developed country like the Netherlands, it is found that only about 36% of the healthcare workers were able to complete all the critical steps [26]. These observations suggest that there is a major gap in skill related to INCS counseling in across the nations, rather than just a local issue. The educational system must be improved to include simulation-based training and mandatory hands-on workshops that allow students and professionals to practice essential steps repeatedly and understand their rationale.

The high proportion of pharmacists demonstrating steps 1,2,4, and 12 correctly (>80%) likely reflects common-sense knowledge (shake, remove dust cap, hold the bottle, replace the cap) that is often taught in basic therapy discussions. However, steps like bending the head forward or cleaning the nozzle were rarely done correctly (<40%). This may cause improper drug disposition, irritation in the throat, and increased risk of contamination [27]. Similarly, only about 10% of participants were counselled about the nozzle being away from the nasal septum, which reduces the risk of nasal irritation, dryness, and epistaxis, and improves drug absorption from the lateral nasal wall [12,27]. In addition, the steps necessary to remove mucus or debris or obstruction from the nose and reduce throat irritation, i.e, blowing the nose before use and exhaling through the mouth, were only performed by about 10% of participants [12]. Patients who are not taught to clean the spray tip may experience clogging or contamination.

These differences align with prior studies indicating that procedural complexity and lack of Continuing Medical Education (CME) or training contribute to inconsistent adherence to medical device protocols [28]. The current study highlights that even pharmacists, who are trained professionals, often lack full mastery of device use and suggests there is a need to improve the curriculum of Pharmacy and improve the landscape of CME in Nepal.

One of the important differences was qualification. BPharm graduates were about 97% less likely to demonstrate incorrect technique than DPharm graduates. The latter finding reflects the differences in Nepal’s educational system. The 3-year Dpharm program in Nepal has traditionally emphasized dispensing skills, whereas the Bpharm and PharmD curricula include more clinical training.

Shrestha et al. found that Nepal’s conventional pharmacy education is mostly lecture-based and industrially oriented, with limited practical training in hospitals [29]. Bhuvan and Subish also documented the challenges in transitioning to PharmD in Nepal with a focus on patient care and pharmaceutical care [30]. This highlights a need for a gradual change in current policy. Medical devices training should be included in the Dpharm degree, and seminars or workshops should be carried out involving Dpharm students and graduates. Pharmacy regulators in Nepal, like the Nepal Pharmacy Council (NPC) or the Department of Drug Administration (DDA), may consider upgrading community pharmacists’ credentials or introducing minimum competency assessments for patient counseling.

Educational aids were also very important in the improvement of INCS counselling. In this study, it was found that pharmacists who used educational materials were much more proficient. This is similar to findings of other studies where pharmacist-led interventions with practical demonstrations and the use of leaflets dramatically improved patient technique in [25,31]. These educational leaflets significantly reduce the cognitive load of pharmacists and ensure the completeness of all steps. These aids also engage patients through teach-back, reinforce learning beyond completeness and boost the pharmacist’s confidence and professionalism.

In our study, both the increasing age (> 26 years old) were significantly associated with improved INCS counselling proficiency. The study conducted in Korea also found that with increased years of experience, the proficiency in patient counselling regarding topical corticosteroids significantly improved [32]. Thus, suggesting increased clinical exposure, trainings, mature communication skills and more frequent patient interaction may contribute to their better proficiency.

Interestingly, participants counselling INCS more than 4 times weekly have a higher proficiency of almost 11 times compared with that of participants counselling occasionally. This relationship likely reflects that a moderate counselling volume provides sufficient repetition to hone skills, confidence while excessive patient load and task interruptions may reduce time for careful demonstration and feedback [33,34]. Simulation training could help low-counseling pharmacists achieve similar proficiency without relying on clinical exposure.

The analysis of this survey revealed that males have about two times higher odds of proficiency than females regarding INCS counselling among the Bpharm graduates. It reflects that there is a difference in the opportunity for workshops and CME or cultural bias. There is a need for the investigation to analyze the barriers associated with the female Bpharm graduates during training or a hands-on workshop.

In our study, most pharmacists (90%) lacked specific training. This recommends that continuing professional development (CPD) for pharmacists in Nepal is sorely needed. According to a recent analysis of CPD in Nepal, continuing pharmacy education (CPE) is still in its infancy; therefore, working pharmacists are not informed of the latest treatments or best practices [35]. Establishing regular INCS technique workshops or integrating device training into the curriculum could narrow the gap. Given pharmacists’ accessibility in rural and urban Nepal [36,37]. Such measures could rapidly propagate correct practice.

The poor skills in pharmacists’ INCS technique are concerning but can be resolved. Targeted training in Nepal could help pharmacists improve their skills quickly. Emphasizing AR and device technique in undergraduate pharmacy programs and requiring competency demonstration during examinations could have a lasting impact. In addition, public health campaigns might encourage patients to ask pharmacists for a demonstration of INCS counselling. In the long term, strengthening pharmacy education and integrating pharmacists into asthma/allergy care pathways will benefit Nepal’s healthcare system by improving primary-level management of chronic respiratory diseases.

## Limitations

This study was conducted in urban Kathmandu, so it may not generalize to rural areas, where pharmacies are fewer and mainly operated by trained dispensers. This study is a cross-sectional design, so it cannot prove causality. Some of the findings may be extreme due to small subgroups or model overfitting. Finally, due to the presence of an interviewer might have influenced the performance, i.e, the Hawthorne effect, possibly inflating technique scores. However, due to the low proficiency observed among the participants, any such effect was limited.

## Conclusion

This study highlights that more than half of the participants don’t have adequate skills to demonstrate a proper intranasal corticosteroid spray usage technique. It is mainly caused due to the poor exposure to this topic in their curriculum during their diploma or graduation classes. In addition, there is very limited training and seminars during their college days or after registration as a pharmacist. Resolving this problem should be one of the important tasks for the Nepal Pharmacy Council and the Health Ministry, as allergic rhinitis is very common in Nepal. Upgrading pharmacy curricula, mandating continuing education, and providing standardized counseling materials may empower pharmacists to counsel correct technique.

## Declarations

### Ethics approval and consent to participate

Ethical approval reference no. 210 (6-11) E2, 080/081 was taken from the Institutional Review Committee of institute of Medicine, Tribhuvan University before the commencement of the study. Informed consent was taken from the participant before any data were collected from the study site.

## Consent for publication

Not Applicable

## Availability of data and materials

The data that support the findings of this study is uploaded in zenodo data repository and the doi link is https://doi.org/10.5281/zenodo.15767865.

## Authors’ contributions

Conceptualization: Dr. Amar Prashad Chaudhary & Suraj Thakur

Methodology: Dr. Amar Prashad Chaudhary & Suraj Thakur

Formal analysis and investigation: Dr. Amar Prashad Chaudhary

Writing – original draft preparation: Dr. Amar Prashad Chaudhary & Suraj Thakur

Writing – review and editing: Dr. Amar Prashad Chaudhary

Supervision: Shiva Kumar Sah

## Data Availability

The data that support the findings of this study is uploaded in zenodo data repository and the doi
link is https://doi.org/10.5281/zenodo.15767865

## List of Abbreviations

AR: Allergic rhinitis
ARIA: Allergic Rhinitis and its Impact on Asthma
Bpharm: Bachelor in Pharmacy
CHAID: Chi square Automatic Interaction Detector
CME: Continuing Medical Education
CPD: continuing professional development
CPE: continuing pharmacy education
DDA: Department of Drug Administration
DPharm: Diploma in Pharmacy
INCS: Intranasal corticosteroid spray
Mpharm: Master in Pharmacy
NPC: Nepal Pharmacy Council
PharmD: Doctor in Pharmacy
SPSS: Statistical Package for Social Sciences
STROBE: Strengthening the Reporting of Observational Studies in Epidemiology

## Acknowledgements

I would like to express my sincere gratitude to my professors, teachers and academic staff of Tribhuvan University for their immense support and guidance. I am also thankful to the pharmacist of Tribhuvan University Teaching Hospital and my beloved friends.

